# Where Adults with Heart Failure Die: Insights from the CDC-WONDER Database

**DOI:** 10.1101/2024.09.17.24313849

**Authors:** Farman Ali, Shaaf Ahmad, Aman Ullah, Adarsh Raja, Faizan Ahmed, Prinka Perswani, Ahsan Alam, Jishanth Mattampuram, Talha Maniya, Hamza Janjua, Tyler J. Bonkowski, Aravinda Nanjundappa

## Abstract

**BACKGROUND:** Heart failure (HF) is associated with high mortality rates and substantial healthcare costs. Although there is increasing emphasis on introducing palliative care for patients with HF, it is not well characterized where adults with HF spend their final days before death.

**STUDY AIM:** This study analyzed the locations and circumstances of death among adults with HF in the United States using data from the CDC-WONDER database.

**METHODOLOGY:** The study examined mortality data of individuals aged ≥ 20 years, with HF listed as the underlying cause of death between 1999 and 2023. The place of death was categorized as the emergency room (ER), hospice/nursing home, inpatient medical facility, or home. Multivariable logistic regression was used to determine the relationship between death location and demographic factors.

**RESULTS:** From 1999 to 2023, HF-related deaths decreased from 1999 (3.60% and 143.6 AAMR) to 2010 (3.47% and 123.1 AAMR). From 2010 onwards, a gradual rise is seen, with the rate of HF deaths reaching 5.18% and 168.1 AAMR in 2023. Notably, deaths at home increased from 18.41% (50,648 of 275,132) in 1999 to 33.47% (132,470 of 395,826) in 2023 and deaths in hospice/nursing homes increased from 30.95% (85,144 of 275,132) in 1999 to 34.71% (116,634 of 336,014) in 2017 and then sudden fall was observed until 2023 to 29.54% (116,931 of 395,826). Older adults (65+) were more likely to die in inpatient facilities. Gender, ethnicity, and urbanization influenced the place of death, with males, White patients, and those residing in large metropolitan areas more likely to die in medical facilities.

**CONCLUSIONS:** This study highlights the changing patterns in the locations of death among HF patients, emphasizing the need for improved home and hospice care services. Addressing disparities in healthcare access and enhancing palliative care are essential for improving end-of-life experiences. Further research is needed to investigate the factors that contribute to these trends.

## INTRODUCTION

Heart failure (HF), owing to its overwhelming global prevalence, was first recognized as an epidemic by Braunwald et al. in 1997.^1^ Since then, substantial leaps in disease management compounded by an aging population have culminated in a massive surge in HF prevalence. Conservative estimates place the current HF prevalence at 64.3 million people worldwide.^2^ But the situation is particularly dire in the United States, where 1 in 4 persons is expected to suffer from HF in their lifetime.^3^ This widespread prevalence of HF becomes particularly concerning when analyzed in the context of the high mortality associated with the syndrome. With some studies approximating 5-year survival rates as abysmally low as 50%, a survival statistic worse than that for most cancers.^4–6^ Given this context, robust management of HF has duly taken center stage in the panorama of cardiovascular disease management, and palliative care has emerged as a cornerstone of optimal disease management strategies. Significant reduction in symptom burden, coupled with decreased acute health care use associated with palliative care intervention in advanced HF, has positioned it favorably, especially when HF-related medical costs are projected to reach $69.8 billion by 2030.^7^ Thus, in recent years, great emphasis has been placed on improving both the quality and uniform accessibility of home and hospice-based healthcare interventions for HF management, as is evident by its inclusion in guidelines for clinical practice.^8^

Following this preface, analyzing HF mortality by place of death becomes particularly relevant when trying to gauge the impact of this shift in HF management strategies to include end-of-life and home/hospice care on mortality trends. Such an analysis can help us ascertain the extent to which this approach has been adopted and its effectiveness in curbing disease burden, in addition to identifying care gaps in comprehensive healthcare delivery for combating heart failure.

## METHODOLOGY

### DATA SOURCES AND EXTRACTION

Data on different causes of death and mortality are accessible in public records from the National Center for Health Statistics (NCHS). Additionally, we utilized aggregated data from the Centers for Disease Control and Prevention’s Wide-ranging Online Data for Epidemiological Research (WONDER) database for the same period. Every death involving heart failure in persons above the age of 20, as indicated by the ICD Code (I50) as the “underlying cause of death,” was covered. The physician’s report on the death certificate identified the “underlying cause of death” as the illness or trauma that set off a sequence of clinical events that finally resulted in death. Excluded from consideration were deaths brought on by suicide, mishaps, murders, self-inflicted wounds, ongoing investigations, or unknown causes of death. According to HHS regulation 45 CFR 46.101(c), informed consent was deemed unnecessary, and there was no need for institutional review because the data were de-identified and accessible to the public.

### VARIABLES

Our variable of interest was the location of death, categorized as an emergency room (ER), a hospice facility or nursing home, an inpatient medical facility, or the decedent’s residence. The age (which was categorized into four ranges: 20-34, 35-49, 50-64, and 65 years), sex (male and female), race (White, Black, American Indian, Asian or Pacific Islander), and ethnicity (non-Hispanic and Hispanic) of the deceased were also analyzed. We also considered the decedent’s geographic identity, which was determined by the county population and categorized using the NCHS rural-urban classification scheme: areas classified as “Large Metropolitan” have a population of one million or more, “Medium Metropolitan” have a population between two and ninety-nine thousand, “Small Metropolitan” have a population between fifty and two hundred thousand, and “Non-Metropolitan or Rural” have a population under fifty thousand. These variables were analyzed to identify mortality patterns across various sociodemographic patterns in the United States.^9^

### STATISTICAL ANALYSIS

Our study aims to explore the relationship between the place of death and sociodemographic factors using multivariable logistic regression across four models. Binary indicators were employed to distinguish between the place of death (1) and other locations (0). The analysis was controlled for age, sex, race, and ethnicity, and odds ratios (OR) and 95% confidence intervals (CI) were calculated. Huber-White patients standard errors were used to evaluate the robustness of the confidence intervals, accounting for unknown correlations in the outcome. P<0.05 was considered statistically significant, and IBM SPSS version 26 (IBM Corp., Armonk, NY, USA) was used for analyses.

## RESULTS

### YEARLY TRENDS

Between 1999 and 2023, a total of 7,644,759 deaths were attributed to HF, with female deaths comprising 53.4% of the total, while deaths accounting for 87.9%, and deaths in adults 65 years of age or above representing 90.3% of total HF deaths. **(Table 1)** provides a summary of the aggregated data for the place of death based on HF decedent characteristics. Over the duration of the study period, there was a significant increase in HF-related deaths. Yearly trend analysis revealed that HF-related mortality decreased from 1999 (3.60% and 143.6 AAMR) to 2010 (3.47% and 123.1 AAMR), with a sudden reversal in 2010 onwards, culminating in HF deaths reaching a high of 5.18% and an AAMR of 168.1 in 2023. (**Figure 1a and Table 1)**. There was also a decrease in the proportion of HF deaths occurring in medical facilities, from 45.13% (124,171 of 275,132) in 1999 to 32.44% (128,412 of 395,826) in 2023. The proportion of HF deaths in emergency rooms has remained approximately the same over this period: 5.51% (15,168 of 275,132) in 1999 and 4.55% (18,013 of 395,826) in 2023. But a significant increase in the proportion of deaths at home, rising from 18.41% (50,648 of 275,132) in 1999 to 33.47% (132,470 of 395,826) in 2023, has been recorded. Throughout the study period, the percentage of fatalities in hospice/nursing institutions increased from 30.95% (85,144 of 275,132) in 1999 to 34.71% in 2017 (116,634 of 336,014) and before diminishing from 34.06% (119,224 of 350,013) in 2018 to 29.54% (116,931 of 395,826) in 2023. **(Table 1)** The distribution of inpatient mortality among the four age groups is depicted in (**Figure 1B**), wherein inpatient deaths make up the largest percentage of deaths across all age groups. Younger age groups recorded higher numbers of death at medical facilities, in contrast to their older counterparts in the 50 years of age and older subset, where the largest percentage of deaths occurred in hospice/nursing homes **(Figure 2)**. was found to be higher in medical inpatient facilities (OR: 0.277; 95% CI, 0.175-0.436) and lower in outpatient ERs (OR: 0.001; 95% CI, 0.0002-0.017), hospice/nursing homes (OR: 0.274; 95% CI, 0.174-0.434), and home (OR: 0.135; 95% CI, 0.066-0.274).

**Table 1.** Aggregated data for adults for places of death by decedent characteristics for Heart Failure (1999-2023)

| **Decedent characteristics** | **Total n (%)** | **Medical facility inpatient (%)** | **Medical facility-outpatient or ER (%)** | **Decedent home (%)** | **Hospice or nursing facility (%)** |
| --- | --- | --- | --- | --- | --- |
| Number of deaths | 7,644,759 (100%) | 2,731,582 (35.7%) | 379,317 (4.96%) | 2,078,986 (27.2%) | 2,454,874 (32.1%) |
| **Age, years** |  |  |  |  |  |
| 20-34 | 20,704 (0.27%) | 11,647 (56.2%) | 3,369 (16.2%) | 4,892 (23.6%) | 796 (3.84%) |
| 35-49 | 110,382 (1.44%) | 54,541 (49.4%) | 17,168 (15.5%) | 32007 (29%) | 6,666 (6.03%) |
| 50-64 | 600,568 (7.85%) | 281,488 (46.8%) | 66,636 (11%) | 182,891 (30.4%) | 69553 (11.6%) |
| 65+ | 6,913,105 (90.4%) | 2,383,906 (34.9%) | 292,144 (4.22%) | 1,859,196 (26.8%) | 2,377,859 (34.4%) |
| **Sex** |  |  |  |  |  |
| Female | 4088199 (53.4%) | 1,366,621 (33.4%) | 172,323 (4.22%) | 1,021,977 (25%) | 1,527,278 (37.4%) |
| Male | 3556560 (46.6%) | 1,364,961 (38.4%) | 206,994 (5.82%) | 1,057,009 (29.7%) | 927,596 (26.1%) |
| **Race** |  |  |  |  |  |
| White | 6,727,596 (87.9%) | 2,326,064 (34.6%) | 292,735 (4.35%) | 1,836,567 (27.3%) | 2,272,230 (33.8%) |
| Black | 749,720 (9.81%) | 331,634 (44.2%) | 74,704 (9.97%) | 190,945 (25.4%) | 152,437 (20.4%) |
| American Indian | 37,059 (0.48%) | 15,527 (41.9%) | 2,713 (7.31%) | 11,625 (31.4%) | 7,194 (19.4%) |
| Asian or Pacific Islander | 130,384 (1.71%) | 58,357 (44.7%) | 9,165 (7.02%) | 39,849 (30.5%) | 23,013 (17.7%) |
| **Hispanic patients patient Origin** |  |  |  |  |  |
| Hispanic patients | 358,353 (4.68%) | 156984 (43.8%) | 23,012 (6.4%) | 113,253 (31.6%) | 65,104 (18.2%) |
| Non-Hispanic patients | 7,286,406 (95.3%) | 2,582,839 (35.4%) | 355,404 (4.9%) | 1,962,784 (26.9%) | 2,385,379 (32.8%) |
| **Urbanization** |  |  |  |  |  |
| Large Metro | 3,493,124 (45.6%) | 1,322,797 (37.9%) | 175,461 (5.03%) | 939,609 (26.9%) | 1,055,257 (30.2%) |
| Medium/ small Metro | 2,516,252 (32.9%) | 851,188 (33.8%) | 118,926 (4.72%) | 700,517 (27.8%) | 845,621 (33.6%) |
| Rural | 1,635,383<br>(21.3%) | 557,597 (34.1%) | 84,930 (5.19%) | 438,860 (26.8%) | 553,996 (33.9%) |

**Figure 1a.**
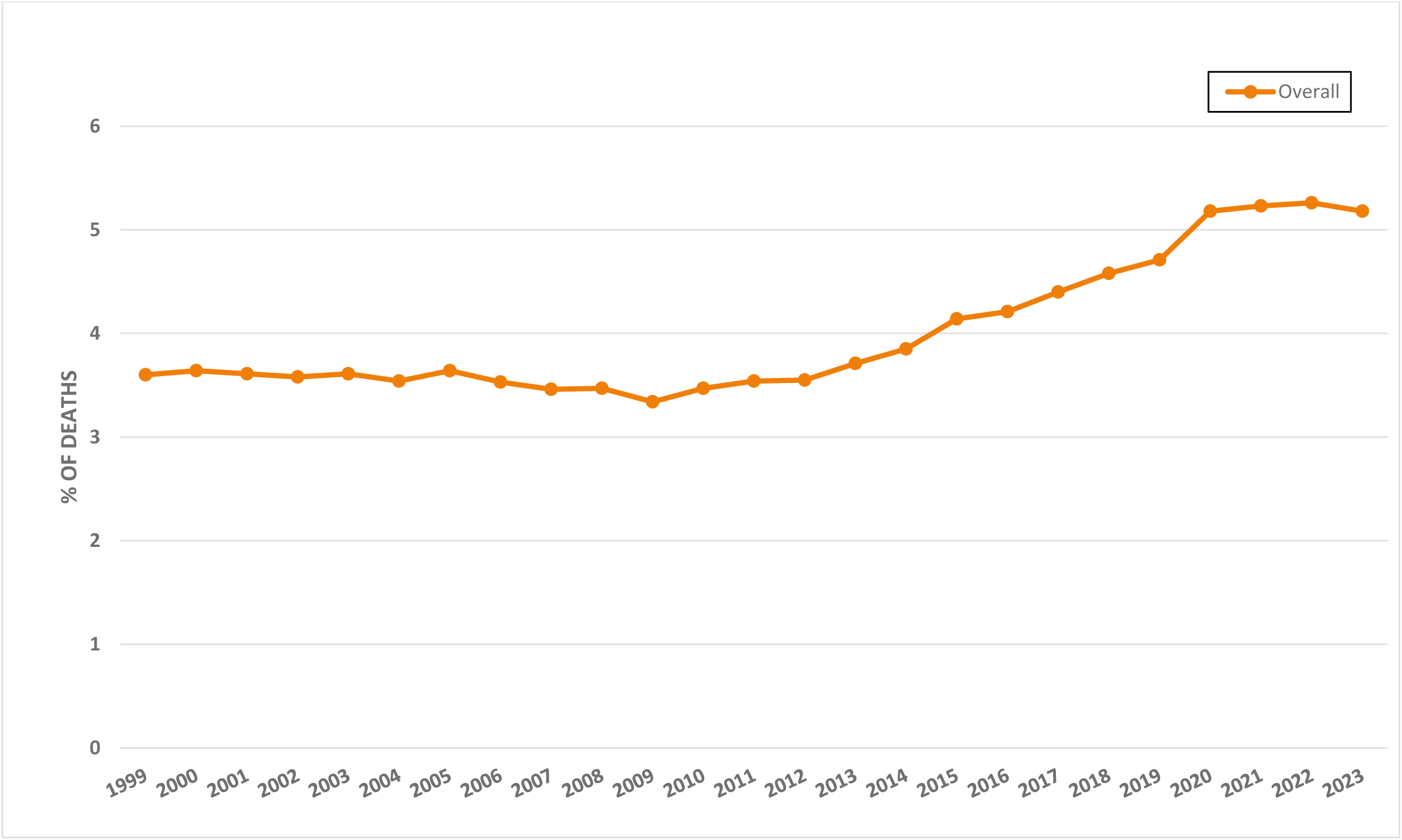
Yearly trends for Heart Failure in Adults (1999-2023).

**Figure 1b.**
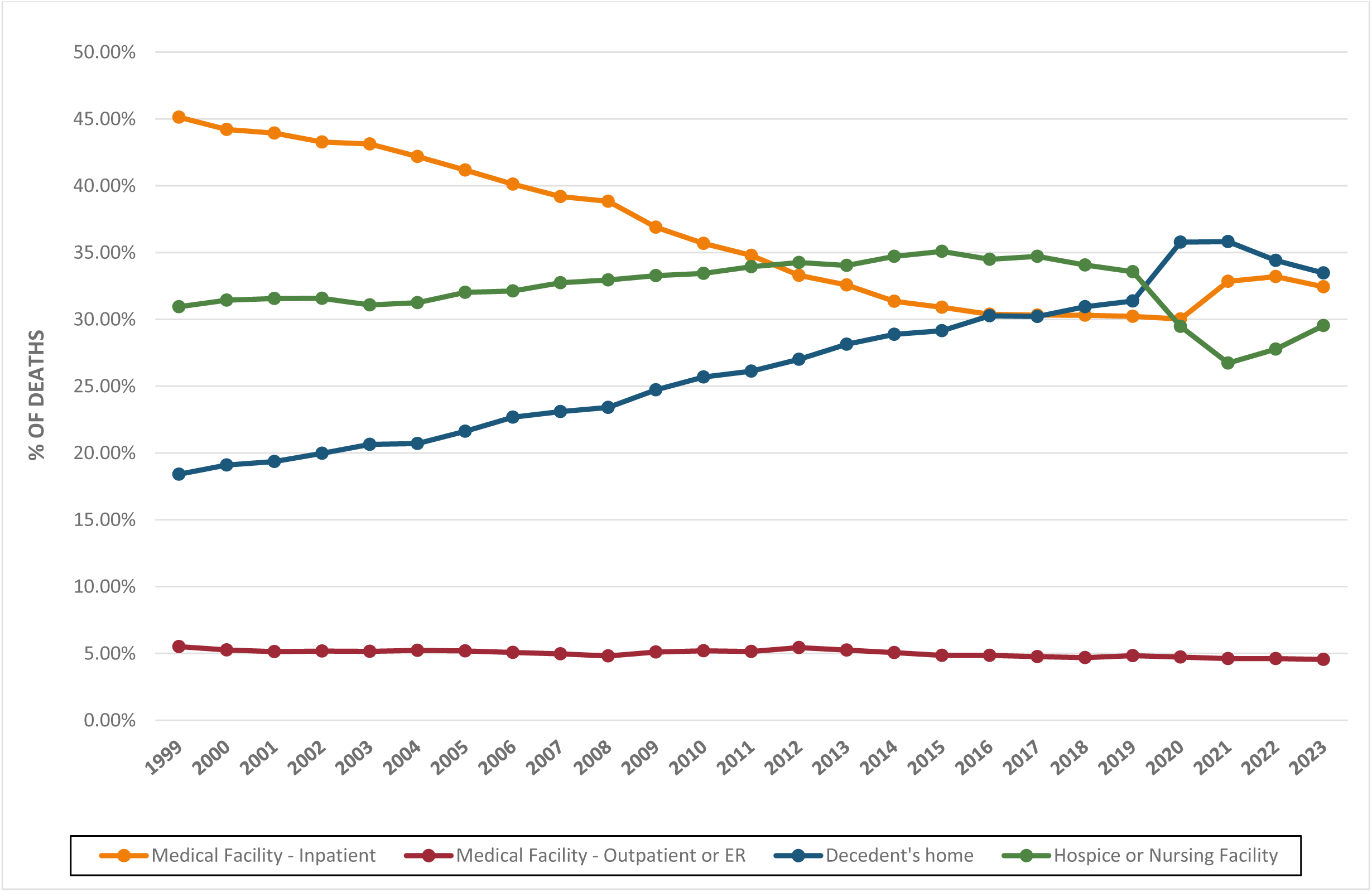
Yearly trends in places of deaths for Heart Failure in Adults (1999-2023).

**Figure 2.**
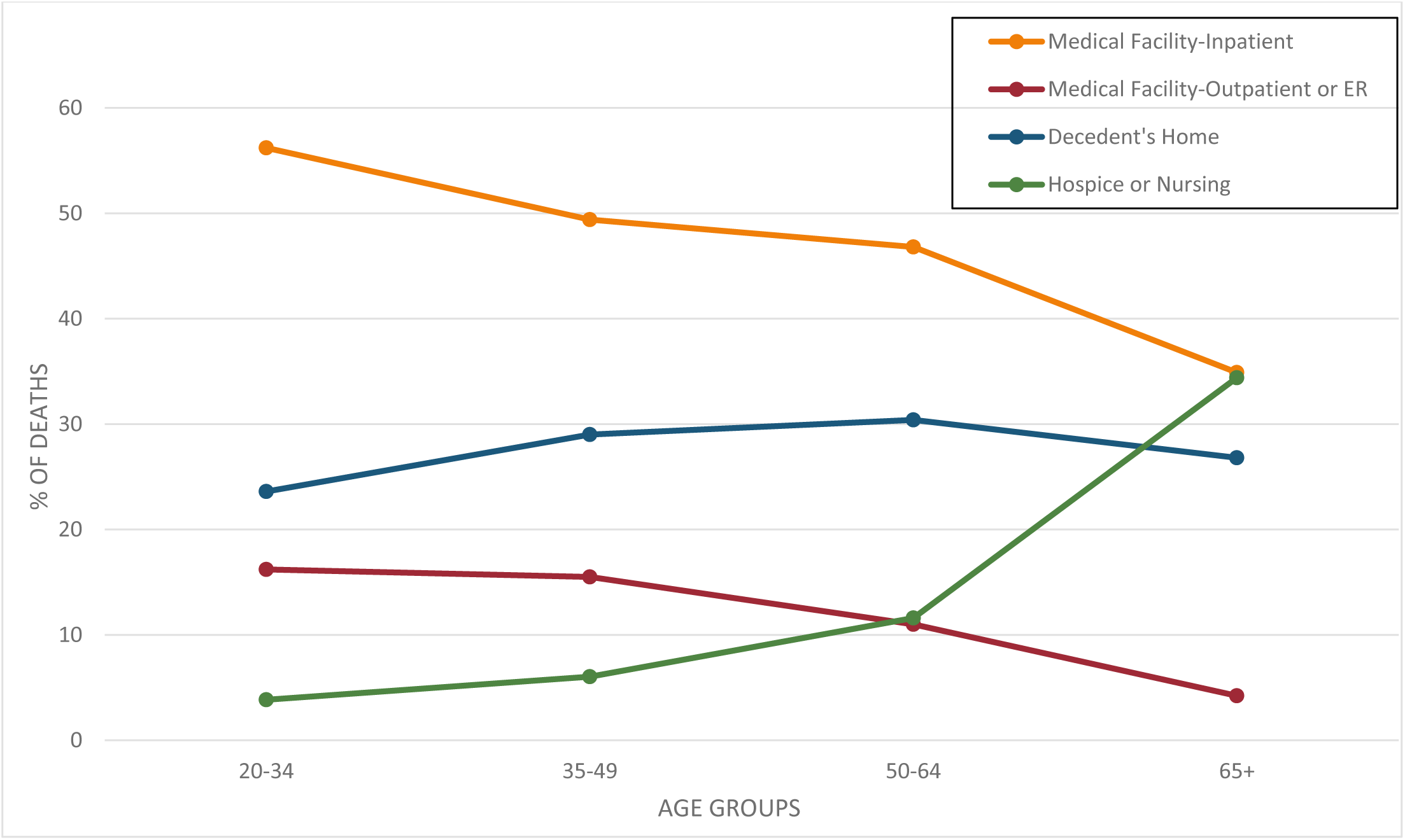
Trends in places of death resulting from Heart Failure in Adults plotted across Age Groups.

### LOGISTIC REGRESSION FINDINGS

The likelihood that a young person with heart failure (20–34 years old) would die in a medical inpatient facility was higher (OR_20-34_: 1.65; 95% CI,1.59-1.72) than that of middle-aged (35–49 years old) people (OR_35-49_: 0.95; 95% CI, 0.94-0.97). The likelihood of dying at a medical inpatient facility was higher for the next age cohort with HF (OR_50-64_: 0.78; 95% CI, 0.71-0.85), and it was the lowest for those above 65+ with HF (OR_65 and above_: 0.28; 95% CI, 0.16-0.44). The likelihood of death for heart failure patients over 65 years of age in the outpatient/ER, hospice/nursing homes, and their homes were (OR_outpatient/ER_: 0.001; 95% CI, 0.0002-0.017 and OR_hospice/nursing home_: 0.27; 95% CI, 0.17-0.43 and OR_home_: 0.14; 95% CI, 0.06-0.27) respectively **(Supplemental Table 1).** Males had higher odds of dying in a medical inpatient facility (OR_males_: 0.39; 95% CI, 0.39-0.39), whereas female decedents were more likely to die in a hospice or nursing home (OR_females_: 0.36; 95% CI, 0.25-0.51) **(Figure 3)**. White patients had lower odds of dying at home (OR_home_: 0.14; 95% CI, 0.14-0.14) than at hospice/nursing home (OR_hospice/nursing home_: 0.26; 95% CI, 0.26-0.26), with the highest likelihood of death being at a medical in-patient facility (OR_in-patient_: 0.28; 95% CI, 0.28-0.28). Asian or PI and non-Hispanic patients populations observed similar trends, where their odds of death increased from at home (OR_Asian or PI patients_: 0.19; 95% CI, 0.19-0.20 and OR_non-Hispanic patients_: 0.14; 95% CI, 0.13-0.14) to in hospice/nursing home (OR_Asian or PI patients_: 0.05; 95% CI, 0.05-0.05 OR_non-Hispanic patients_: 0.24; 95% CI, 0.24-0.24), to becoming the highest for in-patient deaths (OR_Asian or PI patients_: 0.66; 95% CI, 0.65-0.67 and OR_non-Hispanic patients_: 0.30; 95% CI, 0.30-0.30) (**Figure 4a and 4b**).

**Figure 3.**
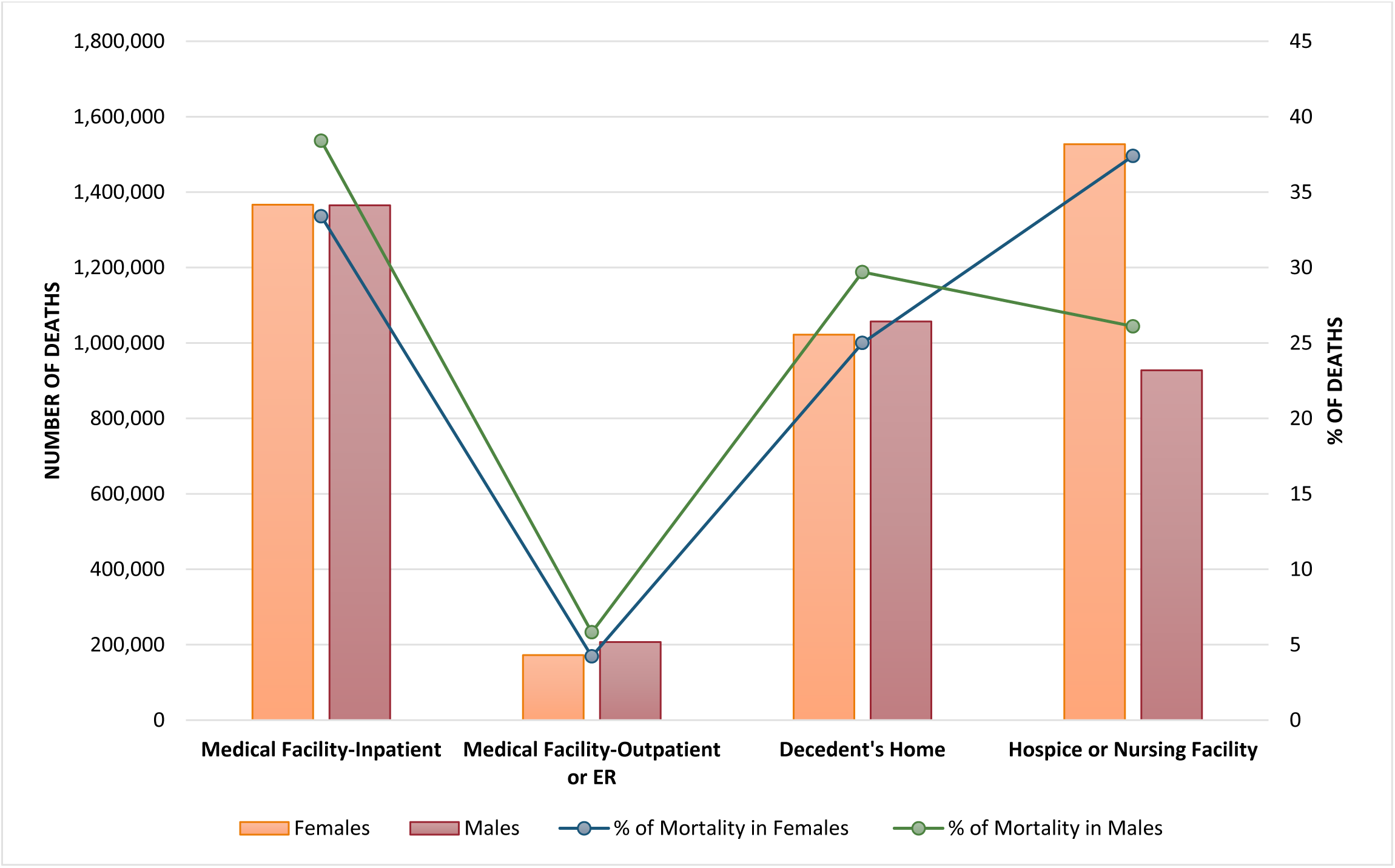
Trends in places of death resulting from Heart Failure in Adults plotted across Sex.

**Figure 4a.**
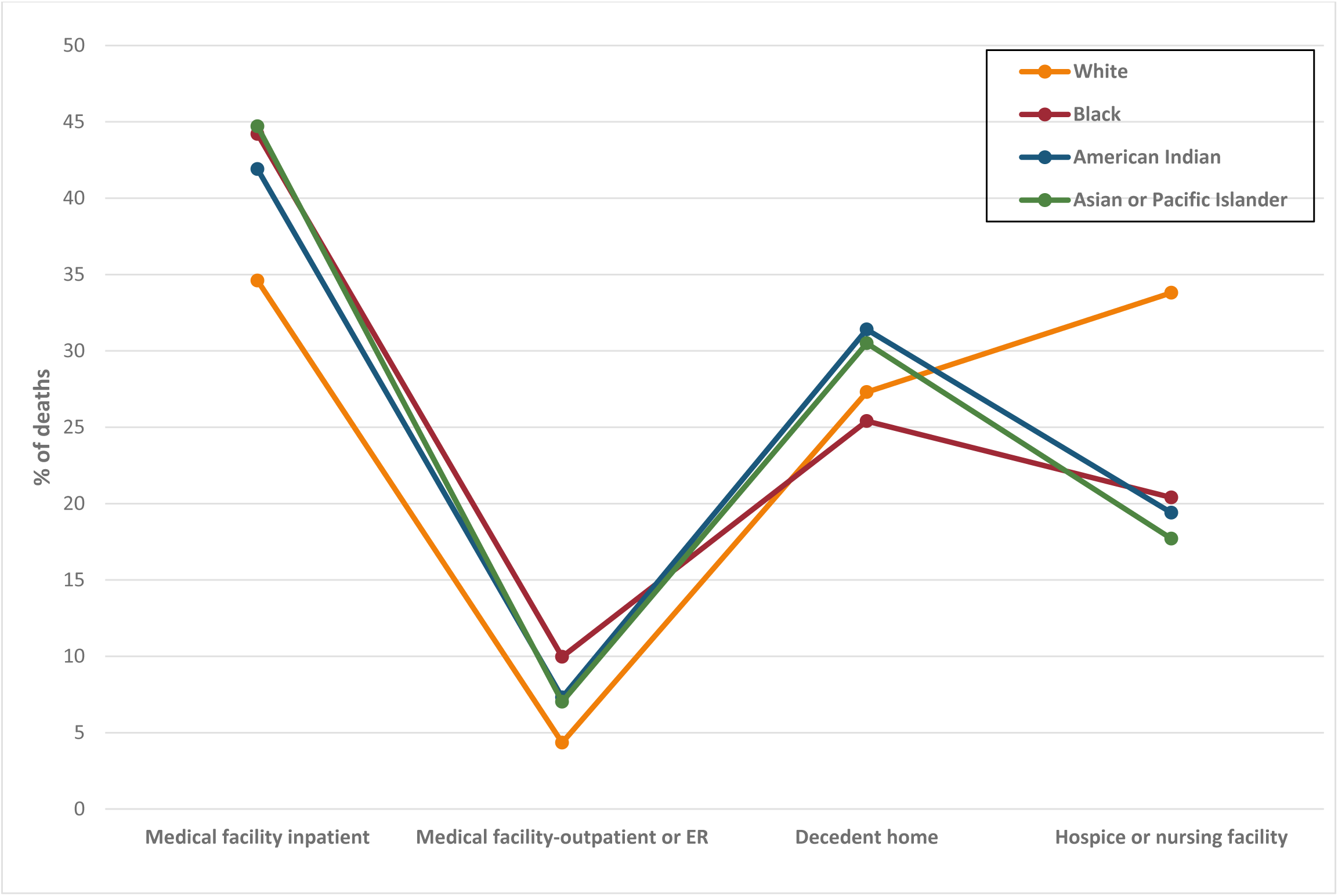
Trends in places of death resulting from Heart Failure in Adults plotted across Race.

**Figure 4b.**
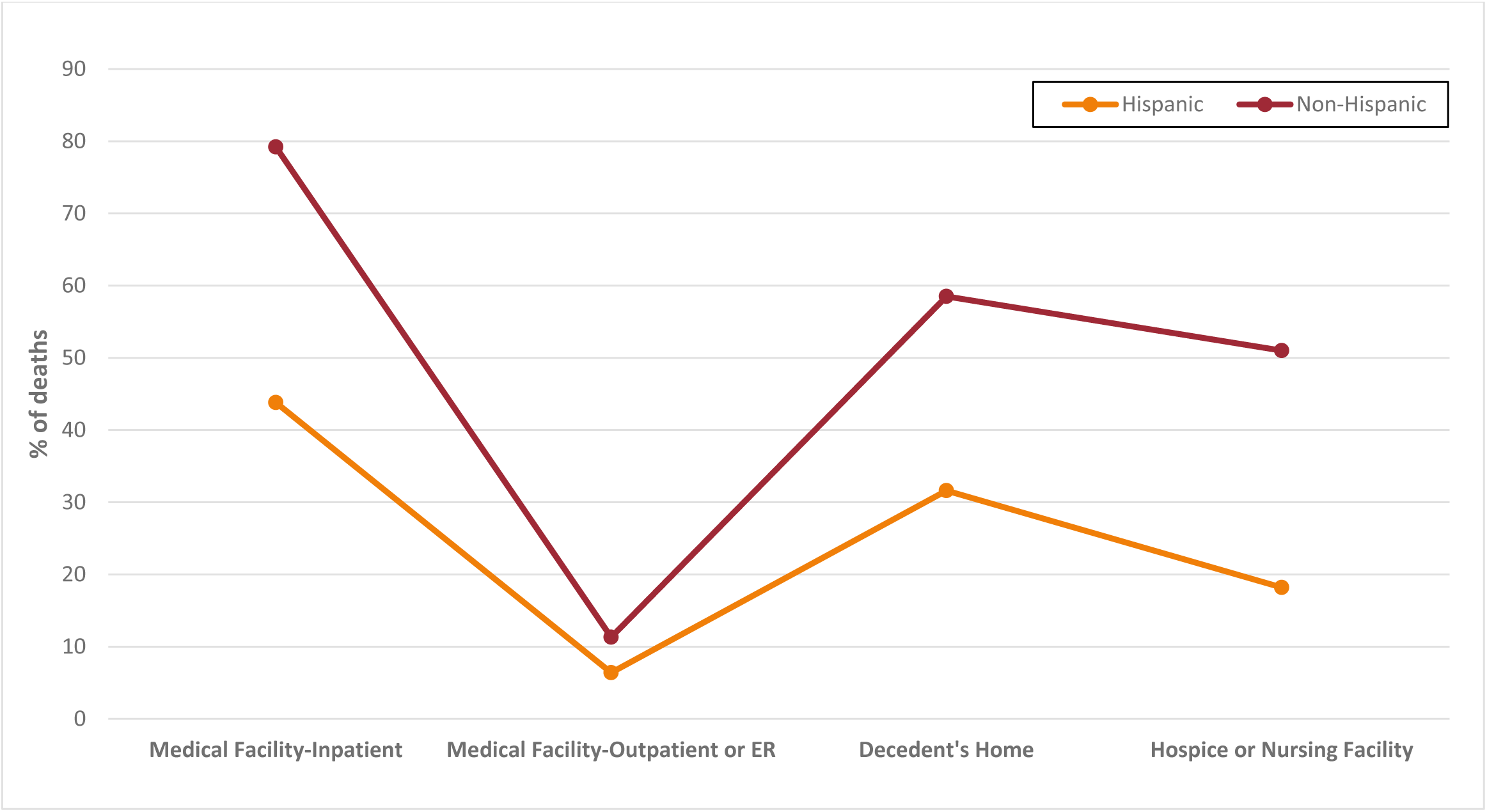
Trends in places of death resulting from Heart Failure in Adults plotted across Hispanic patients patient patients Origin.

Stratification by urbanization revealed that there was a higher incidence of HF mortality in medical inpatient facilities for large metropolitan cities (37.9%, OR: 0.37; 95% CI, 0.37-0.37), followed by medium/small metros (33.8%, OR: 0.26; 95% CI, 0.26-0.26), and finally the rural towns and counties, (34.1%, OR: 0.27; 95% CI, 0.27-0.27) in descending order of likelihood. **(Figure 5)**. The findings of the multivariable logistic regression analysis, which looked at the relationship between the characteristics of the deceased and their place of death, are depicted in Supplemental Table 1.

**Figure 5.**
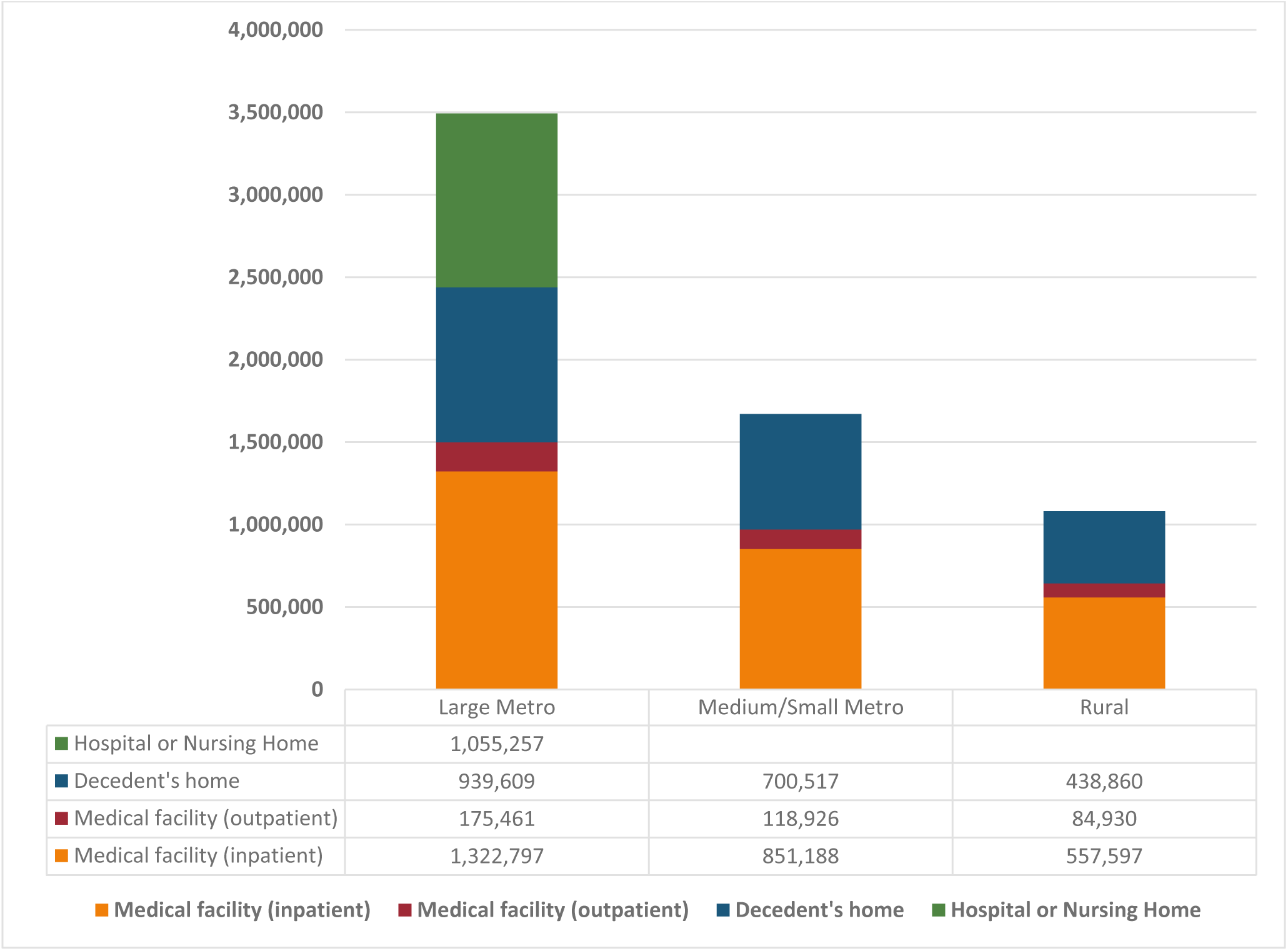
Trends in places of death resulting from Heart Failure in Adults plotted across Urbanization and Age groups.

## DISCUSSION

This is, to the best of our knowledge, the first study to comprehensively evaluate heart failure-(HF) related mortality in the backdrop of ‘place of death’ by utilizing data abstracted from the CDC WONDER database when queried for the years 1999 through 2023. This dramatic reversal in the previously down-trending HF-related deaths^10^ is accounted for, in part, by an increasing prevalence of unmanaged comorbidities, namely, obesity and hypertension. A study by Alpert et al.^11^, exploring the odds of developing congestive HF (CHF) as a function of duration of morbid obesity, noted the probability of developing CHF to be as high as 93% in individuals with >25 years of morbid obesity. The Framingham heart study established two-to three-fold increased risks of developing HF for the hypertensive population. However, managing hypertension in HF has been known to be especially challenging, with outcomes following a J-curve.^12^ Simply put, both an ambitious control of hypertension or a lack thereof are associated with worse HF-related outcomes, necessitating that a balance be struck to optimize survival. This complexity can result in suboptimal disease management, which leads to poorer outcomes in this cohort. A holistic review of HF-related all-cause mortality revealed a remarkable 30% increase for the duration of the study, attributable, in part, to the sudden upturn in mortality rates in 2012 following a steady decline. A finding corroborated by the existing body of literature.^13^ Analysis with the place of death as a co-variate revealed a few key insights. First, even though overall HF-related mortality increased significantly, statistics for deaths encountered in medical facilities, both in and out-patient, observed a downturn from a high of 45.1% in 1999 to 32.4% in 2023. In comparison, statistics for mortality at home reflected a significant increment, from 18.4% in 1999 to 33.5% in 2023-outnumbering the number of HF-related deaths in medical facilities for the first time in history. The trend for deaths in hospice/nursing homes inexplicably peaked in 2017 (34.7%) before diminishing to 29.5% in 2023. Interestingly, deaths in the emergency room hovered over a consistent 5%±0.5% over the past two decades. Second, the percentage of deaths due to HF occurring in medical facilities decreased from 56.2% of all HF-related deaths in 20-34-year-olds to 34.9% for those above 65 years of age. Whereas deaths occurring in the home/hospice/nursing home care settings were the lowest for the 20–34-year-olds (3.8%) while being the highest for the 65 years and older (34.4%). To expand on this further, age-group-based stratification denoted an inverse relationship between increasing age and the odds of dying in hospital. In contrast to this paper that demonstrated higher odds for hospice/nursing home-based deaths in the young,^14^ we showed that the 20-34-years old age stratum had a higher likelihood of in-patient deaths (OR_20-34_=1.65 [1.59-1.72]) in contrast to their older counterparts (OR_65+_=0.27 [0.18-0.44]). As one of the explainers of this disparity, patients younger than 65 with advanced heart failure are more likely to qualify for the receipt of therapeutic interventions such as left ventricular assist devices (LVAD) and heart transplants.^15^ Given their invasive nature and frequent rehospitalizations for intervention-related complications, this subset tends to suffer from greater in-patient mortality than its aged counterpart. Third, grouping outcomes by race revealed that while mortality at home was comparable between the White patients and the Black patients or African American HF populations (27.3% and 25.4%, respectively), the latter had considerably lower hospice/nursing home-related mortality figures in comparison to the White patients (33.8% versus 20.4%). This disproportionately low prevalence of palliative care delivery in the setting of hospice/nursing homes amongst the African American population has been well documented in the existing body of literature.^16^ Their tendency to forgo palliative care services can be attributed, in part, to their religious, spiritual, and cultural beliefs, a general mistrust of the healthcare delivery systems rooted in history, and an under-representation of Black providers in palliative and hospice/nursing home care institutions.^17^ Of note, mortality in hospice/nursing homes was comparable between large metropolitans (37.9%) and the non-core rural cities (34.1%), serving as a potential indicator of effective penetration and acceptance of palliative care services for HF patients in these areas. This is in spite of the dwindling numbers of Medicare-certified hospice/nursing home care centers in these counties, from 871 in 2018 to 824 in 2022.^18^

Multivariate logistic regression analysis revealed that males were more likely to die in-patient (OR_males_=0.39 [0.386-0.389] vs. OR_females_=0.25 [0.15-0.41]) and females in hospice/nursing home (OR_females_=0.36 [0.25-0.51] vs. OR_males_=0.13 [0.12-0.13]). This observation becomes particularly interesting when analyzed in the context of societal traditions and inherent differences in role socialization. A greater prevalence of adherence to the toughness norm amongst men is hypothesized to be a significant barrier to optimal palliative care delivery in a hospice/nursing home setting in this subset.^19,20^ Women, on the other hand, are more likely to prefer end-of-life care, exhibiting greater compliance with palliative care goals.^21^

As depicted above, the rise in hospice /nursing home-based HF-related mortality is multifactorial. An increased awareness of the utility of, and emphasis on, patient-centered end-of-life care might be one of the key facets of this change. Heart failure is not an isolated disease but rather a clinical syndrome causing detriment across a wide spectrum of somatic and psychosocial faculties, resulting in repeated hospitalizations, ultimately culminating in enormous, aggregated costs incurred by the hospitals. Therefore, effective integration of hospice/nursing home-based palliative care was recognized as a potent combat strategy and was incorporated into the consensus statements for managing HF. ^8,22,23^ This shift in strategy might be one of the major catalysts behind the change in place-of-death trend that we underscored in our analysis. It is, however, imperative to mention that while things appear to be changing for the good with an increased utilization of hospice and palliative care services, a lot is still left to be desired. Racial and rural disparities pose the two biggest challenges to optimal and equitable HF management with hospice-based care integration. Educating HF patients about the palliative care available to them, addressing and dispelling misconceptions, facilitating the easy transition of care from the hospital to hospice, and investing in expanding the existing network of Medicare-certified hospice centers into rural towns and counties are the needs of the hour. Advancements in in-patient HF management without an adequate integration of effective home healthcare, hospice, and nursing home care will fail to deliver fitting results. Lastly, robust research into circumventing the roadblocks hampering effective home/hospice-based healthcare is needed to expand the current body of literature and better inform clinical decisions and policy making.

## LIMITATIONS

Limitations of the study include its reliance on death-certificate data abstracted from CDC WONDER, which may not represent the full spectrum of end-of-life experiences for HF patients. The study’s focus on deaths due to HF may overlook individuals with HF who died from other causes, potentially leading to an incomplete understanding of end-of-life care for this population. Additionally, the use of aggregated, de-identified data limits the study’s ability to account for the differences in individual-level clinical characteristics and treatment modalities, which represent crucial determinants in understanding end-of-life care for the HF cohort. This may also obscure essential nuances in the circumstances surrounding deaths in different settings, potentially limiting the depth of insight. Finally, the mortality data from the CDC WONDER database does not touch upon the causation of differentiation in mortality rates in races, gender, urbanization, location of death, and census region.

## CONCLUSION

Our study highlights the evolving patterns in the locations of death among individuals with heart failure from 1999 to 2023. The increase in home and hospice/nursing facility deaths, alongside a decrease in inpatient deaths, underscores the shifting dynamics in end-of-life care preferences and accessibility. Significant factors influencing these trends include age, gender, ethnicity, urbanization, and healthcare policy and technology advancements. Addressing disparities in healthcare access and enhancing palliative care services are critical for improving end-of-life experiences for heart failure patients. Future research should explore the sociodemographic and systemic factors shaping these trends to inform better care practices and policies.

## Data Availability

Data can be provided upon request.

---

**Supplemental Table 1.**
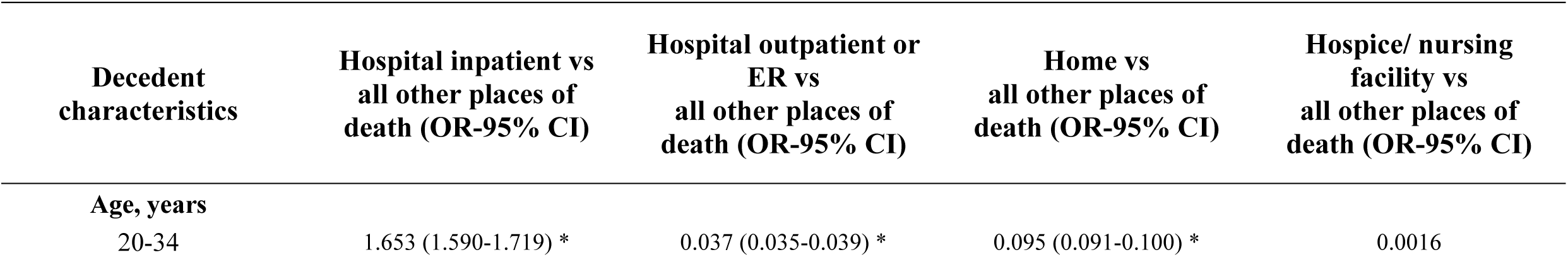
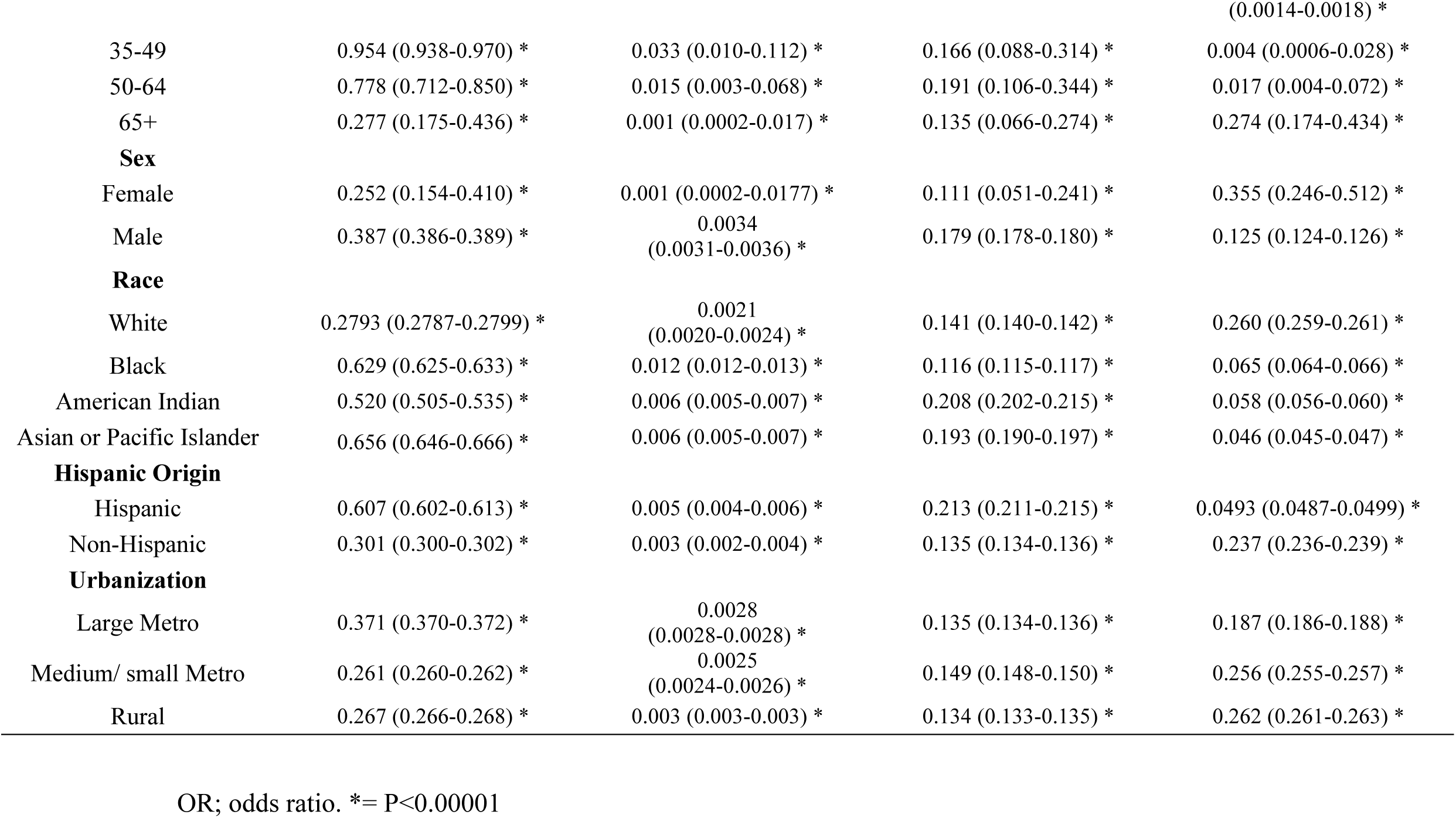
Multivariable logistic regression between decedent characteristics and place of death (1999-2023)

